# A Machine Learning Pipeline for Scalable Annotation of Patient-Ventilator Dyssynchrony from Bedside Ventilator Data

**DOI:** 10.64898/2026.06.11.26355207

**Authors:** Abdulhakim Tlimat, Anoop Mayampurath, Sami Safadi, Jonathan Kalehoff, Nitin Seam, Robert B. Johnson, Sandeep Bodduluri, Surya P. Bhatt, Majid Afshar

## Abstract

**Rationale:** Patient–ventilator dyssynchrony is common in mechanically ventilated patients and associated with worse outcomes, but its detection depends on expert waveform interpretation that is time-consuming, error-prone, and difficult to scale. Machine-learning approaches have been limited by the scarcity of large, expert-labeled waveform datasets, particularly for rare dyssynchrony subtypes.

**Objectives:** To develop and evaluate a scalable annotation pipeline combining semantic-clustering retrieval, expert labeling, and semi-supervised analysis, and to assess whether labels it produces support accurate multi-label classification of dyssynchrony subtypes.

**Methods:** Continuous airway flow and pressure waveforms were acquired from 84 adults receiving invasive mechanical ventilation in two medical intensive care units. Breaths were represented as 512-dimensional vectors, and a semantic interface allowed expert annotators to group morphologically similar breaths s. Two intensivists established reference labels with inter-rater agreement assessed on a 280-breath subset. A multi-label gradient-boosting classifier was trained with decision thresholds tuned on out-of-fold training predictions and evaluated on a held-out test set. A semi-supervised procedure then expanded the labeled set, with performance change assessed against a pre-specified equivalence margin.

**Results:** Of 13,882,390 captured breaths, 1,542,297 were expert-labeled across eight classes and partitioned evenly into training and test sets. Inter-rater agreement yielded a pooled Cohen’s kappa of 0.77. On the held-out test set, the classifier achieved F1 scores from 0.963 to 1.00 across all eight classes, including 0.973 for ineffective triggering and 0.963 for delayed cycling, the two rarest subtypes. Area under the precision–recall curve was at least 0.993 for every class. Semi-supervised expansion after five rounds enlarged the training set from 771,148 to 3,835,056 breaths, with a change in mean per-class F1 of −0.0017 (95% CI, −0.0025 to −0.0009), within the pre-specified equivalence margin of ±0.01.

**Conclusions:** A semantic-clustering annotation pipeline produced a large, expert-labeled dataset of mechanical ventilator waveforms and supported accurate multi-label classification of common and rare dyssynchrony subtypes, providing a scalable foundation for automated dyssynchrony detection.

**Impact Statement:** Detection of patient–ventilator dyssynchrony, including rare subtypes that can generate injurious tidal volumes, has been constrained by the absence of large, expert-labeled waveform datasets. This study shows that a semantic-clustering annotation pipeline can produce an expert-labeled dataset substantially larger than those previously reported and support accurate multi-label classification of common and rare subtypes. Our work provides support for large-scale waveform labeling that is critical for automated dyssynchrony prediction.

## INTRODUCTION

Patient-Ventilator Dyssynchrony (PVD), or ventilator dyssynchrony, occurs when there is a mismatch between the mechanical support provided by a ventilator and the respiratory needs of a patient undergoing invasive mechanical ventilation (IMV).(1–4) The problem of PVD is common, with incidence rates varying between 10% and 85% among critically ill patients receiving IMV.(5) PVD has been associated with harmful tidal volumes and increased transpulmonary pressure, possibly contributing to increased ventilator-associated lung injury (VALI).(6) There is conflicting evidence on whether PVD is associated with worse outcomes in critically ill patients, but patients with increased PVD have higher sedation burden, longer IMV duration, or increased mortality.(7,8)

Monitoring and detecting PVDs is challenging in clinical practice. Methods include visual analysis of ventilator waveforms, but bedside waveform interpretation is error-prone even among experienced clinicians, and achieving competency requires extensive structured training.(9,10) Esophageal pressure measurement and electrical activity of the diaphragm (EAdi) can assist detection, but are invasive and not routinely implemented as standard practice. Diaphragmatic ultrasound offers a noninvasive alternative for identifying patient inspiratory effort but lacks standardization. None of these methods is scalable to continuous automated monitoring across all ventilated patients in an ICU environment.(11)

PVD encompasses multiple subtypes that differ markedly in prevalence.(2,7,8) Ineffective triggering is typically the most common (7,8), others, such as reverse triggering and delayed cycling, occur far less frequently.(12,13) Low prevalence does not imply low importance, as reverse triggering and breath stacking can generate large, uncontrolled tidal volumes and elevated transpulmonary pressures even when infrequent.(5,12–14) Reliable automated recognition of these rare subtypes requires training datasets large enough to contain sufficient examples of each.(2,14) Whether dyssynchrony burden or its detection differs across sociodemographic groups has not been characterized.

Machine learning has been proposed to automate PVD detection, but prior work has been constrained in scope and scale. Two recent narrative reviews characterized the field as limited by small, retrospectively curated datasets, narrow subtype coverage, and offline operation on recorded waveforms rather than continuous bedside capture.(15,16) Across thirteen studies, the cumulative dataset comprised approximately 5.8 million breaths from 332 patients,(15) and Rietveld et al. reported that 84% of reviewed algorithms remained in the development or validation stage, that most operated offline, and that the majority were limited to one or two subtypes, most commonly ineffective triggering and double triggering.(16) Most approaches have also been single-label, classifying each breath as a single subtype and thereby missing the co-occurring events that a clinically meaningful Asynchrony Index requires.(15) In this work, we aim to develop and validate a scalable, end-to-end machine learning pipeline for accurate detection of normal and dyssynchronous breaths from invasive mechanical ventilation in critically ill patients. We hypothesized that a semi-supervised learning approach could extend a smaller set of expert annotations into a large, multi-label annotated waveform dataset spanning all observed PVD subtypes, while preserving labeling accuracy provided by human experts across even the rarest subtypes. This model is intended to support large-scale waveform annotation and quantification of dyssynchrony burden, with clinical application contingent on the patient-level validation that this study does not provide.

## METHODS

### Study Design, Setting, and Patient Population

This is a prospective observational study to collect mechanical ventilator waveform data from patients admitted to two medical intensive care units at the University of Alabama at Birmingham Medical Center between February 15, 2023, and October 5, 2023. The study was approved by the Institutional Review Board at the University of Alabama at Birmingham with a waiver of informed consent given the observational nature of the study (IRB-300010368). All adult patients (age ≥ 18 years) receiving IMV were eligible for inclusion. Data collection began within 24 hours of intubation and continued until one of the following endpoints: extubation, transfer to another unit, transition to tracheostomy, or death. Pregnant patients and prisoners were excluded.

### Ventilator Software Data Capture

Continuous airway flow and airway pressure waveforms were collected at 50 Hz intervals from Puritan Bennett PB980 mechanical ventilators (Medtronic, Dublin, Ireland) using a network of Raspberry Pi edge computing devices. Each Raspberry Pi unit ran internally developed software that interfaced directly with the ventilator to capture breath waveforms in real time. Breath delivery type and mechanical ventilation mode were also recorded from the ventilator at the time of collection. Collected waveforms were streamed to a cloud-based, HIPAA-compliant storage system via Azure IoT Hub (Microsoft, Redmond, WA) in 1-minute batches, without requiring integration with the hospital electronic health record system..

### Breath Segmentation and Waveform Representation

Individual breaths were segmented directly from ventilator output using the breath boundary signals natively provided by the Puritan Bennett PB980 ventilator, which emits a stop signal at the boundary between consecutive breaths. This approach leverages the ventilator’s internal triggering-detection logic and avoids the need for post hoc algorithmic segmentation of the raw waveform. Only breaths delivered during Assist-Control (AC) ventilation, including AC/VC, AC/PC, and AC/VC+ breath delivery types, were retained for analysis.

Each segmented breath consists of simultaneous, variable-length recordings of airway flow and pressure. To produce a fixed-length representation suitable for machine learning, we identified the cycling point, the transition between the inspiratory and expiratory phases, as the first negative flow value in each breath, excluding the initial 100 milliseconds in patient-triggered breaths to avoid spurious detections. The inspiratory segment and expiratory segment were each independently resampled to 128 time points using linear interpolation, yielding a 256-point representation for each signal. The flow and pressure vectors were concatenated to produce a fixed-length feature vector of 512 dimensions per breath. Cycling-point alignment was chosen to ensure consistent positioning of both the inspiratory and expiratory phases across breaths of varying duration. Segmentation and pre-processing were applied identically to all breaths, without reference to patient characteristics.

### Reference Labels by Expert Human Reviewers

Five dyssynchrony subtypes were defined through clinical expert consensus and distinguished from normal ventilation. These five dyssynchrony classes, along with a class denoting normal breath, and two classes for breath delivery type (pressure control vs volume control) yield eight classes in total. Breath delivery type was assigned from the ventilator-reported mode at the time of acquisition rather than inferred from waveform morphology. Because VC+ breaths are delivered as pressure-controlled breaths with an adjusted pressure target, their single-breath flow and pressure morphology are indistinguishable from AC/PC; VC+ and PC breaths were therefore grouped under a single pressure-controlled delivery label. Label definitions and annotation rule sets were established a priori by three intensivists with 5-10 years of post-fellowship experience in mechanical ventilation and patient-ventilator asynchrony. The five subtypes were ineffective triggering (a patient effort that fails to trigger a mechanical breath), flow starvation (delivered flow insufficient to meet inspiratory demand), premature cycling (patient effort continuing after the ventilator cycles to expiration), delayed cycling (patient expiratory effort beginning before the ventilator cycles), and breath-stacking dyssynchrony (two ventilator breaths delivered in rapid succession with insufficient intervening expiratory time). A sixth quality-control category, “Unclassifiable,” was applied to breaths in which signal artifact, atypical morphology, or absence of a morphologically coherent cluster precluded confident assignment. These breaths were excluded from all analyses. Complete definitions and the full waveform criteria for each category are provided in Supplementary Table S1.

To assist human annotation, a custom web-based interface was developed to facilitate large-scale labeling of the breath dataset. Breath waveforms were stored as 512-dimensional vectors in PostgreSQL with the pgvector extension, enabling approximate nearest-neighbor search by cosine similarity within the database. An unlabeled breath was presented as a waveform visualization displaying flow, pressure, and volume signals. The annotator specified the number of nearest neighbors to retrieve, and the system queried pgvector using cosine similarity on the 512-dimensional vector to return that many morphologically similar breaths from the unlabeled pool. The annotator reviewed the resulting cluster and assigned one or more labels to the entire cluster according to the ruleset in Table S1. Setting the neighbor count allowed the annotator to maintain morphological coherence, retrieving fewer neighbors for atypical waveforms and more for common ones.

All initial annotations were performed by a single pulmonary and critical care physician with 2 years of experience in ventilator waveform interpretation. A randomly selected subset of 280 breaths was independently relabeled by a second pulmonary and critical care physician with 5 years of experience, blinded to the original annotations. Inter-rater agreement was quantified using Cohen’s kappa, calculated per label and pooled across all labels. Kappa values were interpreted according to the criteria of Landis and Koch, where values of 0.61–0.80 indicate substantial agreement and values above 0.80 indicate almost perfect agreement.

### Model Development for Supervised Machine Learning

The dataset of labeled breaths was partitioned into a training/development set and a held-out test set using a 50/50 random split stratified by label combination. Partitioning was performed at the breath level by design. Because breaths from the same patient may therefore appear in both partitions, the reported metrics quantify labeling fidelity and are not patient-level generalization estimates. Because each breath can carry multiple labels, we trained one independent binary gradient boosted classifier per label using LightGBM with the scikit-learn MultiOutputClassifier wrapper. Class imbalance was addressed by setting class_weight=‘balanced’ in each binary classifier, which scales the loss contribution of positive examples by the inverse class frequency.

Hyperparameters were tuned by randomized search restricted to the training and development partition, with the held-out test set excluded from all tuning. Twenty-five configurations were evaluated on a 150,000-breath development subsample by macro-averaged area under the precision-recall curve (AUPR) under 3-fold cross-validation, averaged across labels with at least one positive example per fold. AUPR was selected as the tuning criterion because it is threshold-independent and robust to severe class imbalance. The full search grid and the retained configuration are reported in Table S2. For each label, the decision threshold maximizing F1 was selected from out-of-fold training predictions by searching the interval [0.10, 0.95] in increments of 0.01. Thresholds were then held fixed across all subsequent evaluations, including every semi-supervised round.

### Semi-Supervised Learning

To extend labeling to the unlabeled breaths, we employed an iterative semi-supervised strategy that used the model’s own predictions to expand the training set. The unlabeled pool was shuffled prior to the first round to prevent ordering bias. In each round, a 5% chunk of the total unlabeled dataset was drawn sequentially without replacement, and the current classifier assigned a label to each breath. A model-assigned label was retained only when the predicted probability exceeded 0.97, a threshold placed well above the decision boundary given the separation between true-positive and true-negative predicted probabilities in the supervised model. Breaths receiving at least one confident model-assigned label were added to the training set, and all classifiers were retrained from scratch on the expanded set, which combined the original expert labels with the accumulated model-assigned labels.

This process was repeated for up to 20 rounds with early stopping triggered by any of the following criteria: (1) F1 score of any rare class label decreased by more than 0.02 relative to the supervised baseline, (2) no labels were assigned for a rare class label in three consecutive rounds, or (3) macro-averaged F1 across all labels changed by less than 0.001 over four consecutive rounds. All stopping thresholds were specified a priori. Their rationale is described in the online supplement.

### Analysis Plan

Model performance was evaluated on the held-out test set after the supervised baseline and after each semi-supervised round using per-label AUROC, precision, recall, AUPR, and F1 score, as well as micro-averaged, macro-averaged, weighted-averaged, and sample-averaged metrics. The F1 score, the harmonic mean of precision and recall, was the primary performance metric because it summarizes both error types and, unlike accuracy, is not inflated by the large majority of true-negative breaths present for each rare subtype. Analyses were implemented in Python 3.12.11. Software versions and training hardware are listed in the online supplement.

Model performance was not stratified by patient, and heterogeneity across patients was not quantified. All waveforms were acquired from a single ventilator platform in medical intensive care units, so no variation across ventilator model or unit type was available to assess. Model performance was not stratified by sociodemographic groups. The model operates on waveform morphology without access to patient characteristics, and subgroup analysis was not prespecified. No formal sample size calculation was performed. All eligible patients with successful waveform capture during the enrollment window were included, yielding over 1,000 held-out test positives for the rarest class.

## RESULTS

### Study Population

Of the patients receiving invasive mechanical ventilation in the two participating units during the study period, 84 had waveforms captured and analyzed. Capture was limited by the availability of 12 acquisition devices rather than by eligibility. The cohort had a median age of 55.5 years [IQR 43.0–67.0], with 44 females (52.4%) and 40 males (47.6%). Most patients were admitted through the emergency department (75.0%). The cohort had a high severity of illness, with a mean SOFA score of 8.1 (SD 1.9) and in-hospital mortality of 39.3%. Full cohort characteristics are presented in Table 1. No data were missing for the variables reported in Table 1.

**Table 1.** Patient demographics and clinical characteristics (N = 84). Abbreviations: RASS, Richmond Agitation–Sedation Scale; SOFA, Sequential Organ Failure Assessment.

| Characteristic |  | Overall (n = 84) |
| --- | --- | --- |
| Age, median [Q1, Q3] |  | 55.5 [43.0, 67.0] |
| Sex, n (%) | Female | 44 (52.4) |
|  | Male | 40 (47.6) |
| Race, n (%) | Black or African American | 37 (44.0) |
|  | White | 37 (44.0) |
|  | Hispanic or Latino | 4 (4.8) |
|  | Decline/refuse | 4 (4.8) |
|  | Multiple | 2 (2.4) |
| Admission source, n (%) | Emergency department | 63 (75.0) |
| RASS score, mean (SD) |  | −1.8 (1.8) |
| SOFA score, mean (SD) |  | 8.1 (1.9) |
| In-hospital mortality, n (%) |  | 33 (39.3) |

### Breath Annotation

A total of 13,882,390 breaths were collected across two medical ICUs over the study period, representing approximately 9,387 hours of invasive mechanical ventilation time. Volume control (VC) accounted for the majority of ventilation time at 7,289 hours (77.6%), followed by volume control plus (VC+) at 1,220 hours (13.0%), and pressure control (PC) at 879 hours (9.4%). Of the breaths presented for annotation, 1,542,297 received a dyssynchrony or normal label through the pgvector semantic clustering system, distributed across 2,720 clusters with a mean cluster size of 575 breaths. A further 0.13% carried the “Unclassifiable” label and were excluded from downstream analyses. The class distribution of the final labeled dataset is shown in **Table 2**.

**Table 2.** Class distribution of the expert-labeled dataset (N = 1,542,297 breaths). Percentages are the proportion of all labeled breaths carrying each label; because breaths may carry multiple labels (delivery type, and normal versus dyssynchrony), columns do not sum to 100%.

| Class | Number of breaths (% of labeled dataset) |
| --- | --- |
| Normal | 1,383,860 (89.73%) |
| Volume-controlled ventilation | 1,145,112 (74.25%) |
| Pressure-controlled ventilation | 396,776 (25.73%) |
| Flow starvation | 131,988 (8.56%) |
| Premature cycling | 87,784 (5.69%) |
| Breath stacking | 35,688 (2.31%) |
| Delayed cycling | 3,620 (0.23%) |
| Ineffective triggering | 2,048 (0.13%) |
| “Unclassifiable” (excluded from analysis) | 2,071 (0.13%) |

VC+ breaths are included within the pressure-controlled ventilation class, as VC+ is delivered as a pressure-controlled breath and is not separable from AC/PC on single-breath waveform morphology.

Inter-rater reliability, assessed on the 280-breath subset, yielded an overall Cohen’s kappa of 0.77 pooled across all labels, indicating strong agreement between human reviewers. Per-label kappa values are presented in **Table 3** and ranged from 0.13 for delayed cycling to 0.81 for normal breaths. Agreement was substantial to almost perfect for the higher-prevalence classes (normal, 0.81; volume-controlled ventilation, 0.75; pressure-controlled ventilation, 0.75; flow starvation, 0.71) and moderate for the rarer dyssynchrony subtypes (ineffective triggering, 0.66; premature cycling, 0.65; breath stacking, 0.54). Delayed cycling demonstrated the lowest agreement (0.13), reflecting the small number of events identified by the second reviewer (n=2) and the subtlety of its waveform signature.

**Table 3.** Inter-rater agreement between the original annotator (Annotator 1) and a second independent expert reviewer (Annotator 2) on a randomly selected subset of 280 breaths. Cohen’s kappa was calculated per label, with counts reflecting the number of positive labels assigned by each reviewer. The overall Cohen’s kappa pooled across all labels was 0.77.

| Label | Annotator 1<br>positive count | Annotator 2<br>positive count | Cohen's kappa |
| --- | --- | --- | --- |
| Normal | 115 | 107 | 0.81 |
| Volume-controlled ventilation | 208 | 230 | 0.75 |
| Pressure-controlled ventilation | 53 | 36 | 0.75 |
| Flow starvation | 113 | 104 | 0.71 |
| Ineffective triggering | 2 | 4 | 0.66 |
| Premature cycling | 44 | 70 | 0.65 |
| Breath stacking | 36 | 68 | 0.54 |
| Delayed cycling | 27 | 2 | 0.13 |
| **Overall (pooled)** | — | — | **0.77** |

### Supervised Classifier Performance

At decision thresholds tuned on out-of-fold training predictions, the supervised classifier performed strongly across all eight classes (F1 0.963–1.00). Performance was near-perfect for the high-prevalence classes: normal ventilation, volume-controlled ventilation, and pressure-controlled ventilation each reached an F1 of 1.00. The moderate-prevalence dyssynchrony subtypes also performed well, with F1 scores of 0.996 for breath stacking, 0.992 for premature cycling, and 0.990 for flow starvation. The two rarest subtypes achieved F1 scores of 0.973 for ineffective triggering and 0.963 for delayed cycling, despite only 1,023 and 1,810 test-set positives, respectively, indicating that class-weighted training preserved performance even under severe class imbalance. Area under the precision–recall curve exceeded 0.993 for every class (macro AUPR 0.998).

Probability distributions for true positive and true negative predictions were well separated for all eight classes, with output probabilities clustering near 1.0 for true positives and near 0.0 for true negatives across all labels (see Supplementary Figure 01 for an example of every subtype). This separation is consistent with the high-confidence gate applied during semi-supervised expansion.

### Semi-Supervised Learning

Iterative labeling was applied over five rounds before the early-stopping criteria were triggered. The training set expanded from 771,148 breaths at baseline to 3,835,056 breaths at round 5, with 90.3% to 96.0% of candidate breaths accepted per round under the confidence gate. Table 4 shows the F1 progression for each class and macro-averaged F1 across all rounds. Performance on the rare dyssynchrony subtypes remained stable across expansion: ineffective triggering F1 moved from 0.973 at baseline to 0.969 at round 5, and delayed cycling F1 from 0.963 to 0.958. Macro-averaged F1 changed from 0.989 at baseline to 0.987 at round 5, at which point expansion halted on the macro-F1 plateau criterion (change < 0.001 over four consecutive rounds). Across the eight classes, mean per-class F1 differed by −0.0017 (95% CI, −0.0025 to −0.0009) between the supervised baseline and the final semi-supervised model. The two one-sided tests procedure supported equivalence against the pre-specified margin of ±0.01 (90% CI, −0.0024 to −0.0011).

**Table 4.** F1 progression for the two rarest subtypes (ineffective triggering, delayed cycling) and macro-averaged F1 across semi-supervised rounds. Expansion halted after round 5 on the macro-F1 plateau criterion (change < 0.001 over four consecutive rounds).

| Round | Training size | Ineffective triggering F1 | Delayed cycling F1 | Macro F1 |
| --- | --- | --- | --- | --- |
| Baseline | 771,148 | 0.973 | 0.963 | 0.989 |
| 1 | 1,357,670 | 0.971 | 0.963 | 0.989 |
| 2 | 1,969,729 | 0.971 | 0.961 | 0.988 |
| 3 | 2,589,079 | 0.970 | 0.961 | 0.988 |
| 4 | 3,211,433 | 0.970 | 0.961 | 0.988 |
| 5 | 3,835,056 | 0.969 | 0.958 | 0.987 |

Macro-F1 difference, supervised baseline vs. round 5: -0.0017 (95% CI -0.0025 to -0.0009; 2,000 bootstrap resamples of the held-out test set), within the pre-specified $\pm 0.01$ equivalence margin. *F1 values are computed on the held-out test set at the frozen out-of-fold decision thresholds. Training size reflects expert-labeled breaths plus accumulated model-assigned labels.*

## DISCUSSION

Using a semantic annotation interface, we assembled 1,542,297 expert-labeled ventilator breaths spanning all observed PVD subtypes. A multi-label classifier trained on these labels performed strongly across all eight classes, including the two rarest subtypes (F1 0.963 to 1.00), and semi-supervised expansion extended labeling to 3,835,056 breaths with performance equivalent to the supervised baseline within a pre-specified margin.

The 1,542,297 expert-annotated breaths analyzed here exceed the cumulative expert-labeled breath count reported across the thirteen studies in the prior literature.(15,16) These data were captured prospectively and continuously: edge computing devices streamed waveforms directly from bedside ventilators to HIPAA-compliant cloud storage, independent of the electronic health record, rather than relying on the retrospective, vendor-specific session exports that have rate-limited prior datasets. The labeling framework is also multi-label, assigning all co-occurring dyssynchrony subtypes within a single breath, in contrast to the single-label architectures that dominate prior work and that systematically miss co-occurring events.

The central enabler of this dataset was the annotation strategy. Expert breath-by-breath review, the standard approach in prior work, scales poorly to the volume of breaths generated in a modern ICU and has confined previous datasets to tens or hundreds of thousands of breaths. By propagating a single expert labeling decision across a cluster of morphologically similar breaths, the semantic-clustering interface substantially reduced annotation burden while preserving label consistency within clusters, yielding 1,542,297 expert-labeled breaths.

This work builds on prior efforts to quantify dyssynchrony burden through a reported Asynchrony Index,(8,17), which requires complete enumeration of co-occurring dyssynchrony events within each breath. Single-label models systematically undercount co-occurring subtypes and bias the index downward. Among prior approaches, only a small number have attempted broad subtype coverage. Ang et al. evaluated a tri-input convolutional neural network classifying seven subtypes, but on a retrospective dataset and with the explicit caveat that more diverse training data are needed before broader adoption. The multi-label design used here, applied to a prospectively captured dataset spanning all observed subtypes, is structurally suited to the index that downstream clinical tools will require.(18)

Semi-supervised expansion has not, to our knowledge, been applied to ventilator waveform classification, and its behavior here is instructive. A roughly fivefold increase in training set size produced no meaningful change in performance on any class, including the two rarest subtypes, and expansion halted on the convergence criterion rather than on any performance decrement. This suggests that the expert-labeled set already contained nearly all of the learnable signal for these subtypes and that additional model-assigned breaths offered diminishing returns at this scale. Whether this plateau holds beyond the present cohort, ventilator model, and subtype distribution is unknown and would require external datasets to establish.

Several limitations warrant consideration. First, the train/test split was performed at the breath level to evaluate annotation fidelity rather than patient-level generalization; accordingly, the reported metrics should not be read as evidence of generalization to unseen patients. Patient-level generalization would require partitioning at the patient level and has not been established here. Second, the pipeline detects breath-stacking dyssynchrony as a single category but does not resolve its two underlying mechanisms, double triggering and reverse triggering, which share a nearly identical flow and pressure morphology within a single breath; distinguishing them requires information beyond the individual breath, including the temporal pattern and patient effort. Third, agreement between reviewers was poor for delayed cycling (kappa 0.13), driven by the second reviewer identifying two events where the first identified 27. The classification performance reported for this class therefore reflects fidelity to a single annotator’s labels rather than to a consensus reference standard, and should be interpreted accordingly. Fourth, inverse-frequency class weighting was applied without subsequent recalibration, so predicted probabilities are not calibrated and should be treated as ranking scores rather than event probabilities.

## Conclusion

We developed a scalable pipeline for automated annotation of patient-ventilator dyssynchrony, producing a large, expert-annotated mechanical ventilator waveform dataset and a multi-label classifier that performed strongly across all subtypes. The dataset, annotation methodology, and classification framework together provide a reproducible foundation for facilitating patient-level dyssynchrony detection and future work in developing the clinical decision support tools.

**Figure 1.**
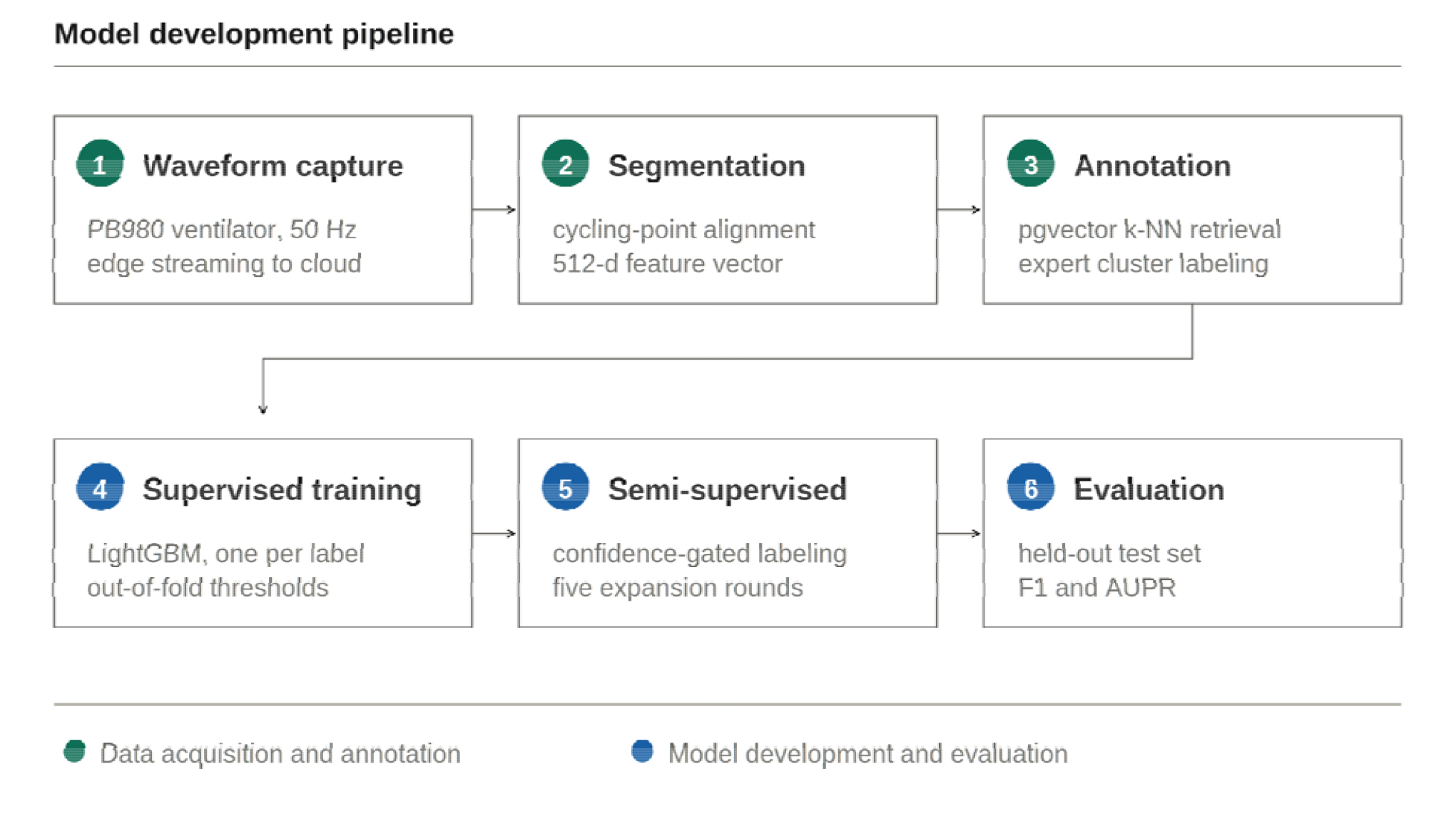
Illustration of steps in model development

**Figure 2.**
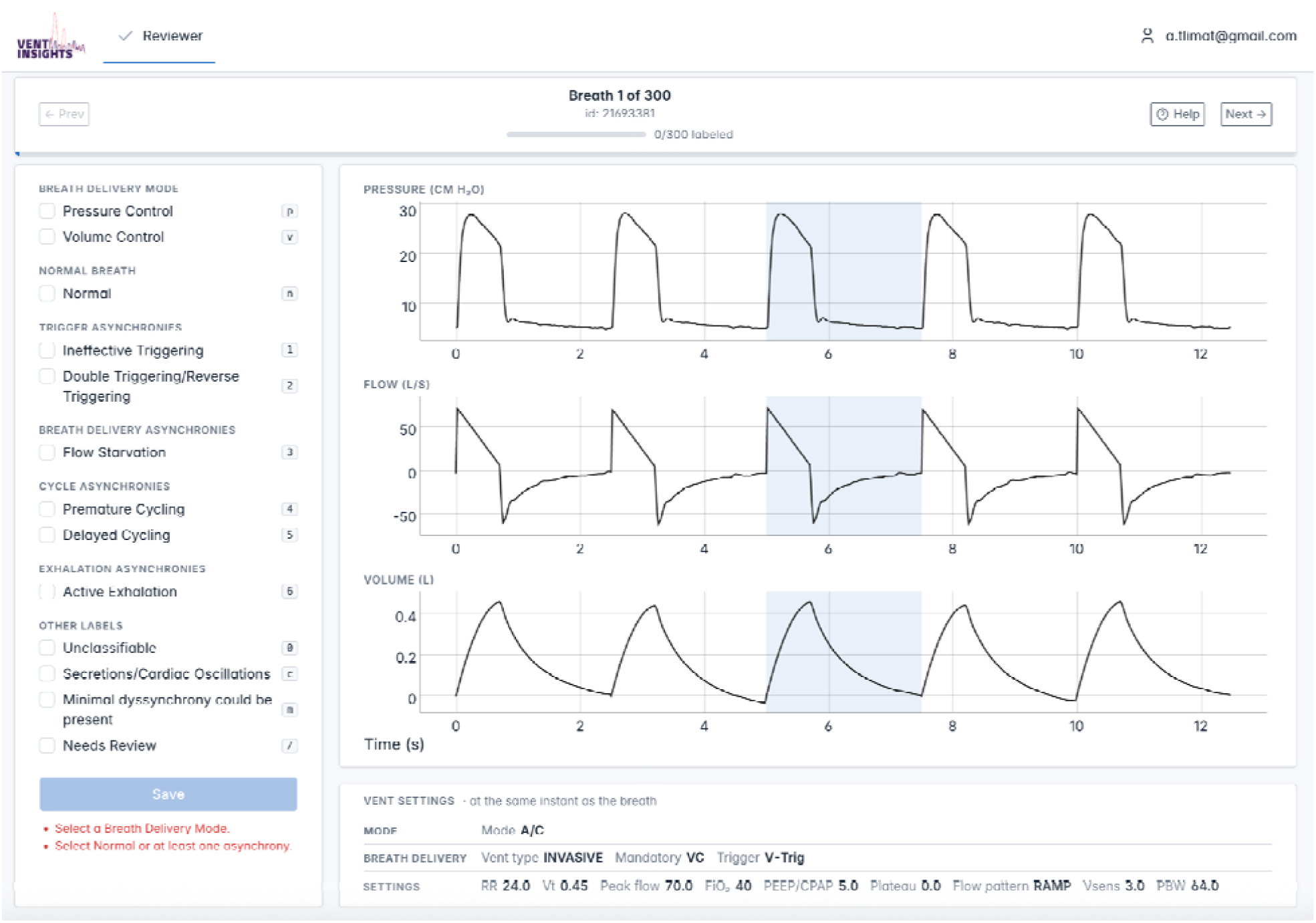
Screenshot of the breath labeling software

## Data Availability

All data produced in the present study are available upon reasonable request to the authors

