## Supplementary Figure S1 for "A Machine Learning Pipeline for Scalable Annotation of Patient-Ventilator Dyssynchrony from Bedside Ventilator Data": Supplementary_Figure_S1.docx

**Supplementary Figure S1.** Representative flow–time and pressure–time waveforms for each annotated dyssynchrony subtype. For flow starvation, premature cycling, delayed cycling, and breath stacking, the waveform is shown separately under volume-control (VC) and pressure-control (PC) delivery; ineffective triggering is shown once, as its expiratory-phase signature is mode-invariant. In each panel, the diagnostic waveform feature specified in the annotation criteria (Supplementary Table S1) is indicated. Each panel shows a single breath drawn from the expert-labeled dataset.

| **Subtype** | **Volume control (VC)** | **Pressure control (PC)** |
| --- | --- | --- |
| **Ineffective triggering** | 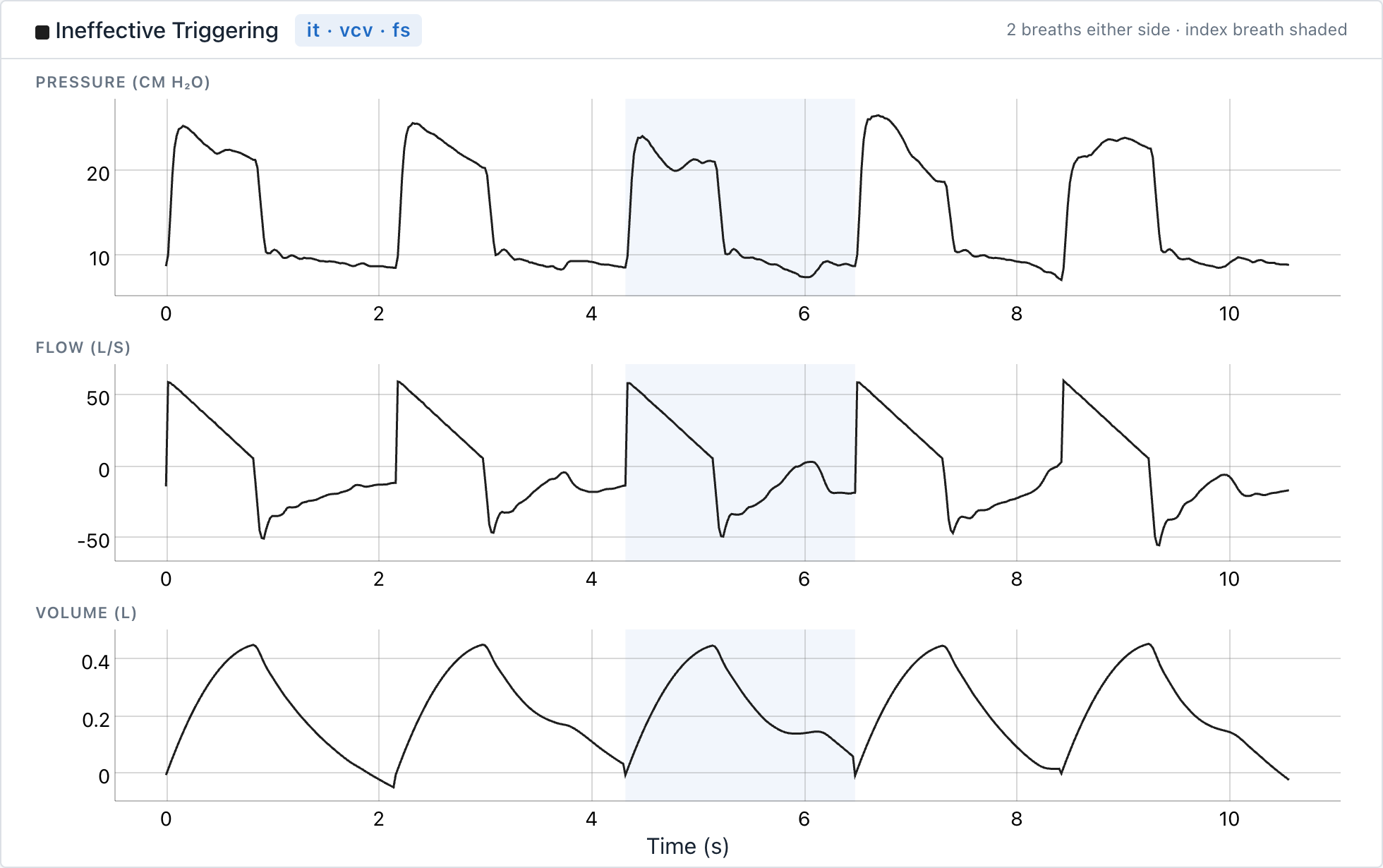 | |
| **Flow starvation** | 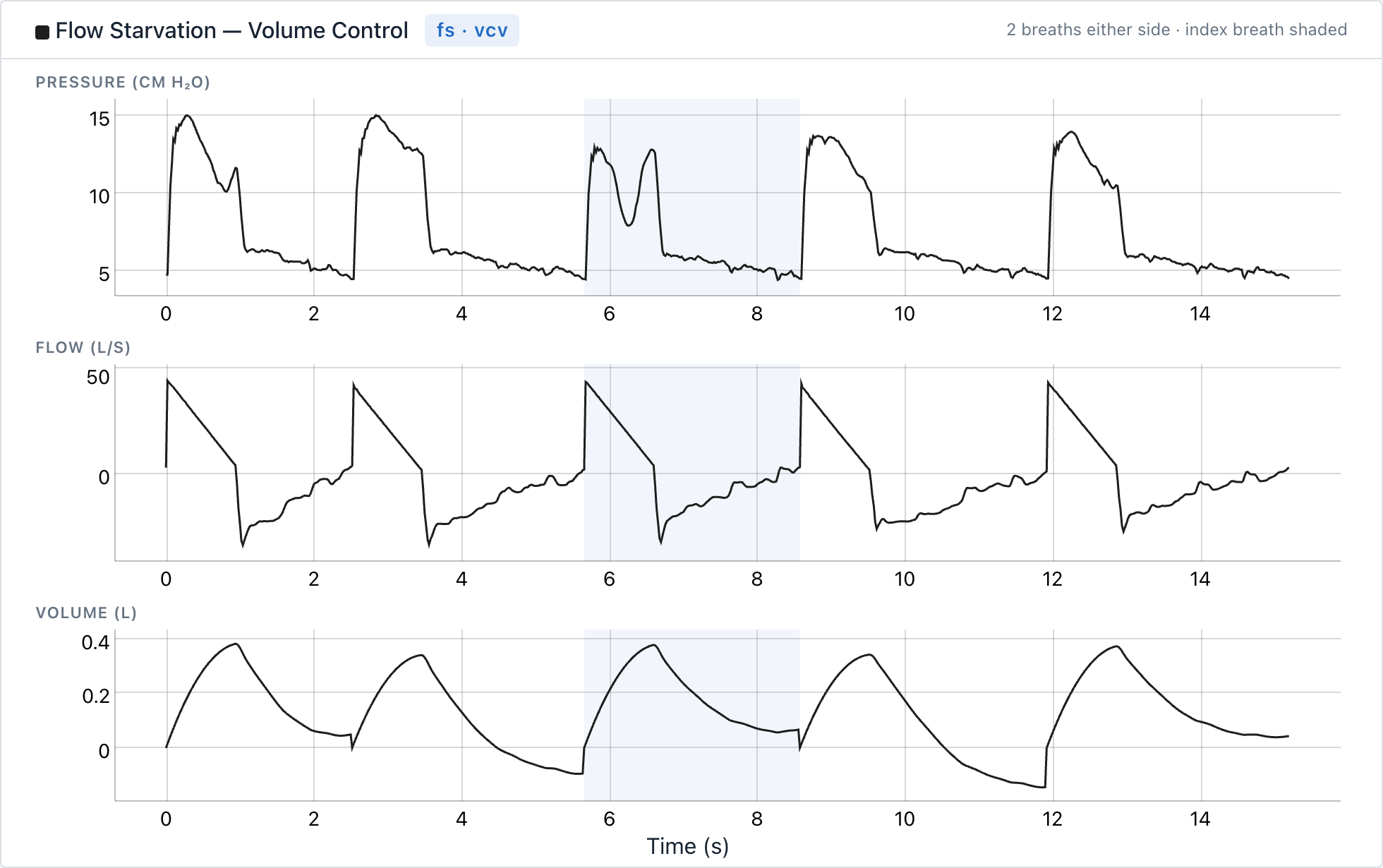 | 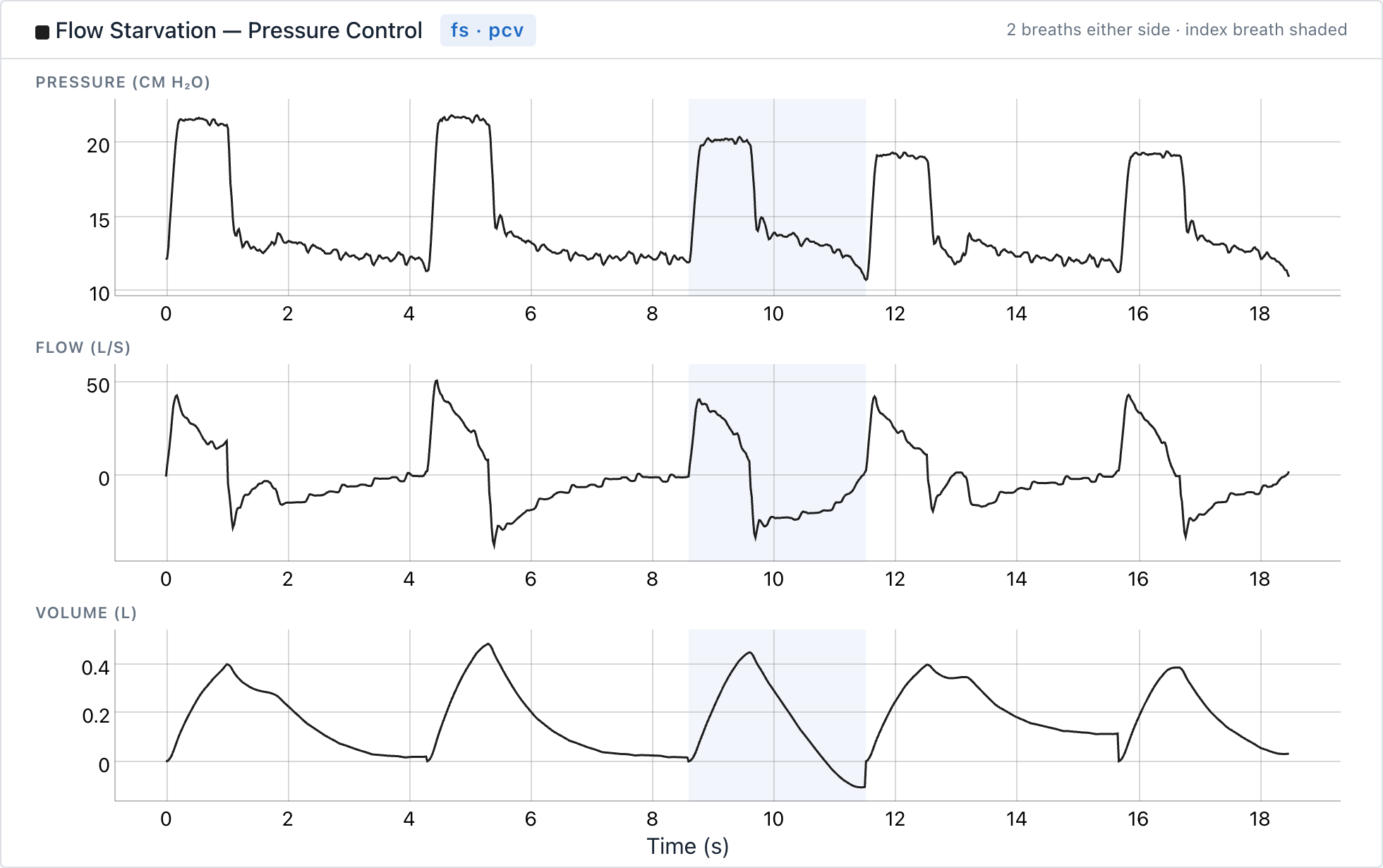 |
| **Premature cycling** | 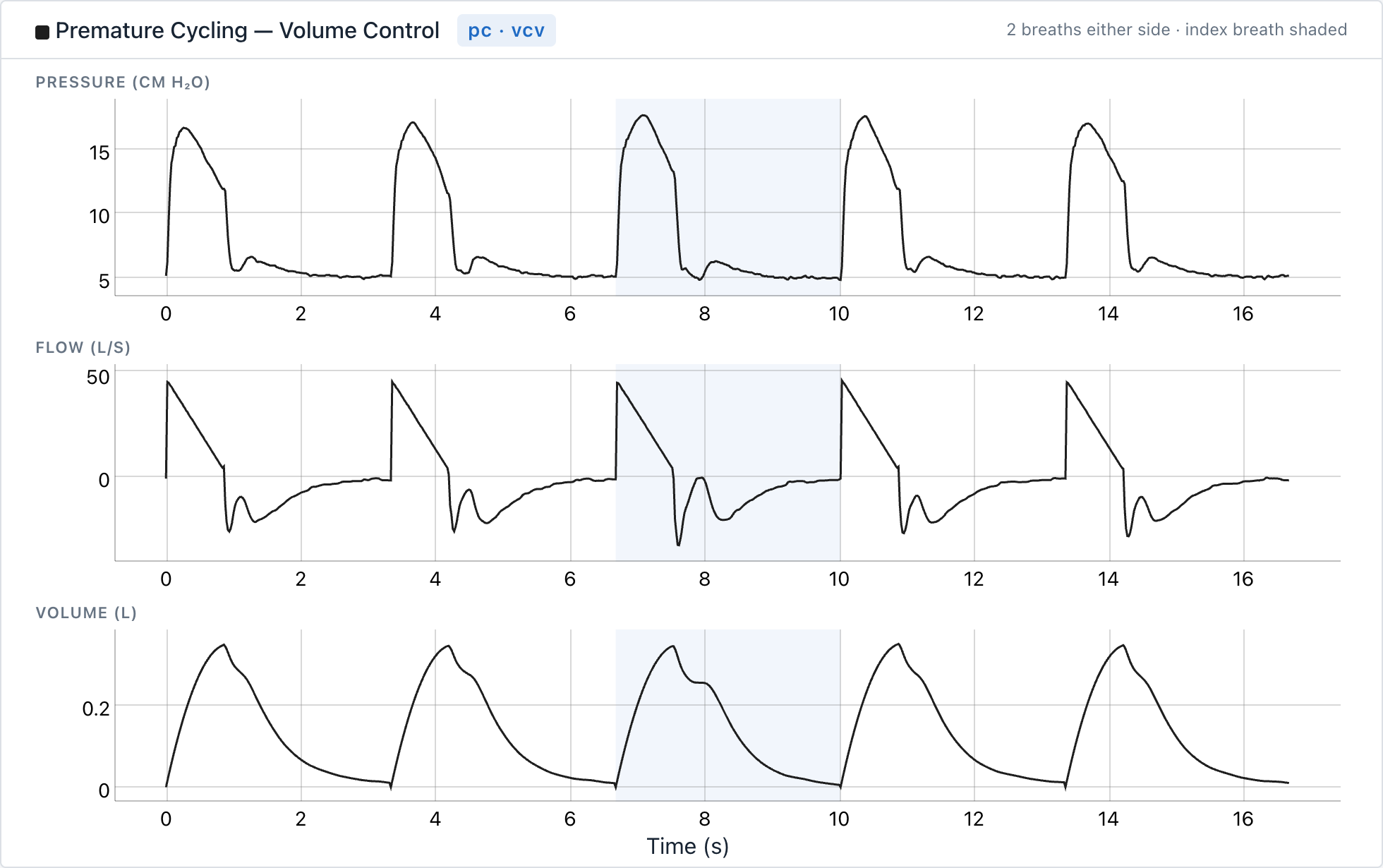 | 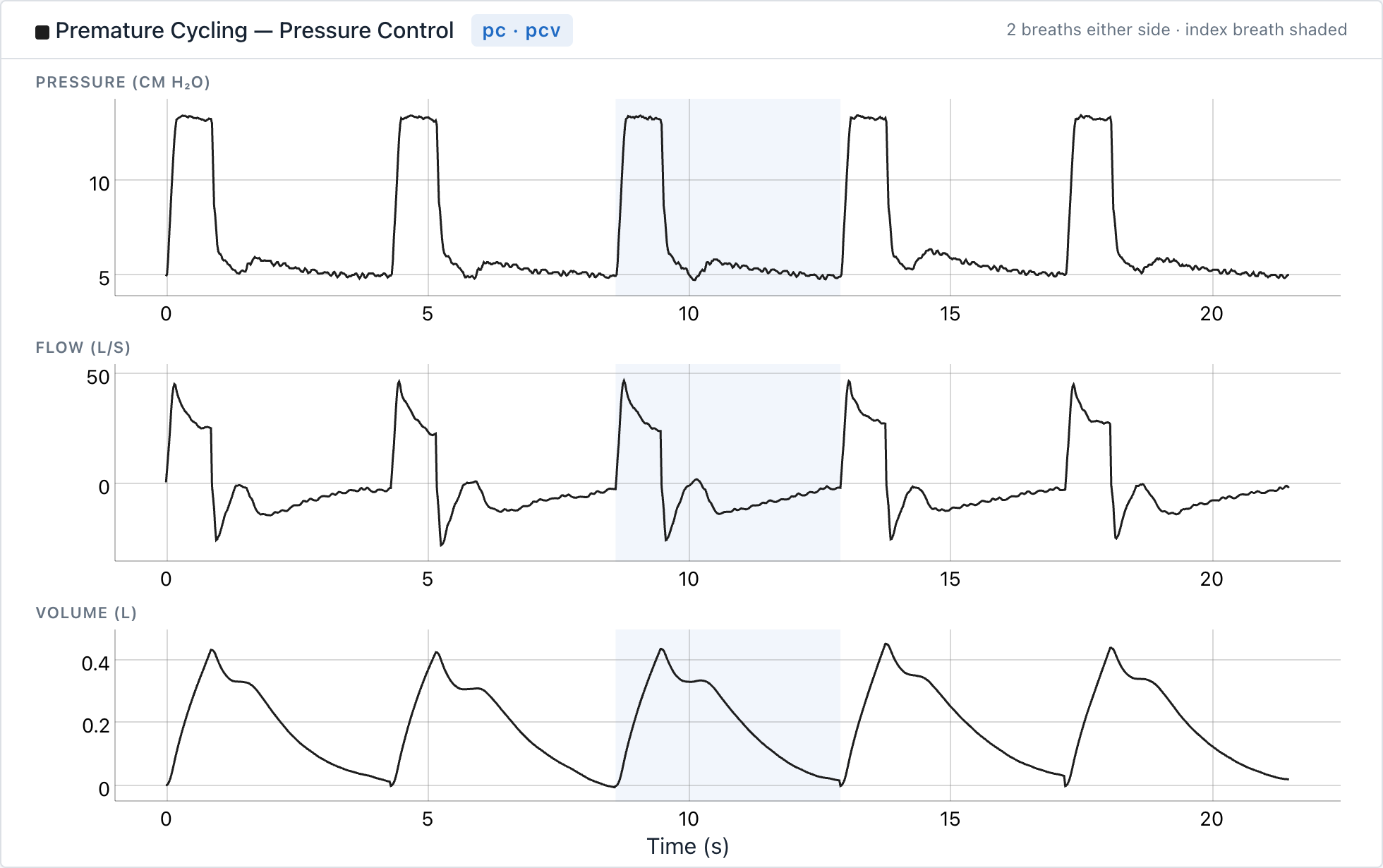 |
| **Delayed cycling** | 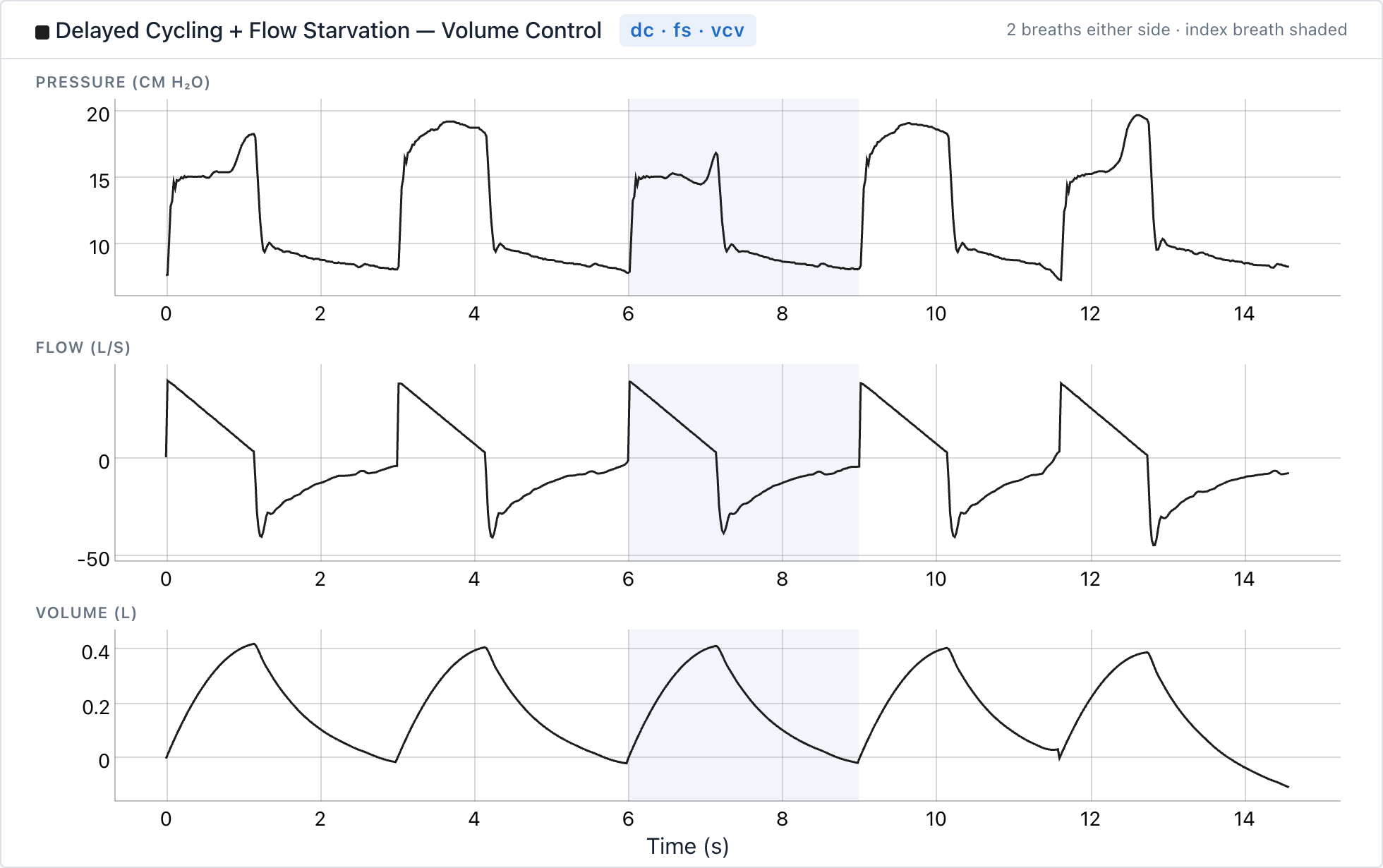 | 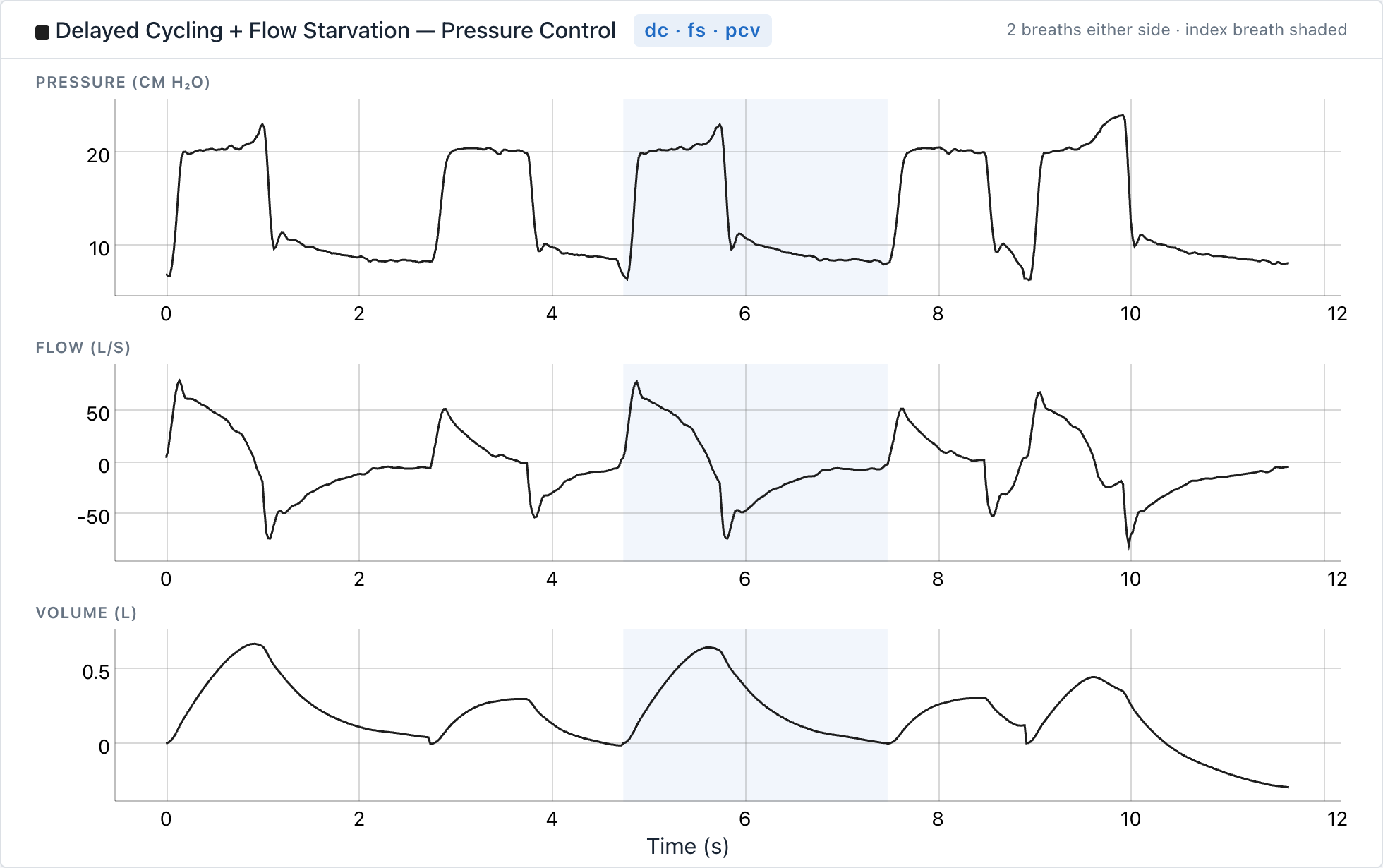 |
| **Breath stacking** | 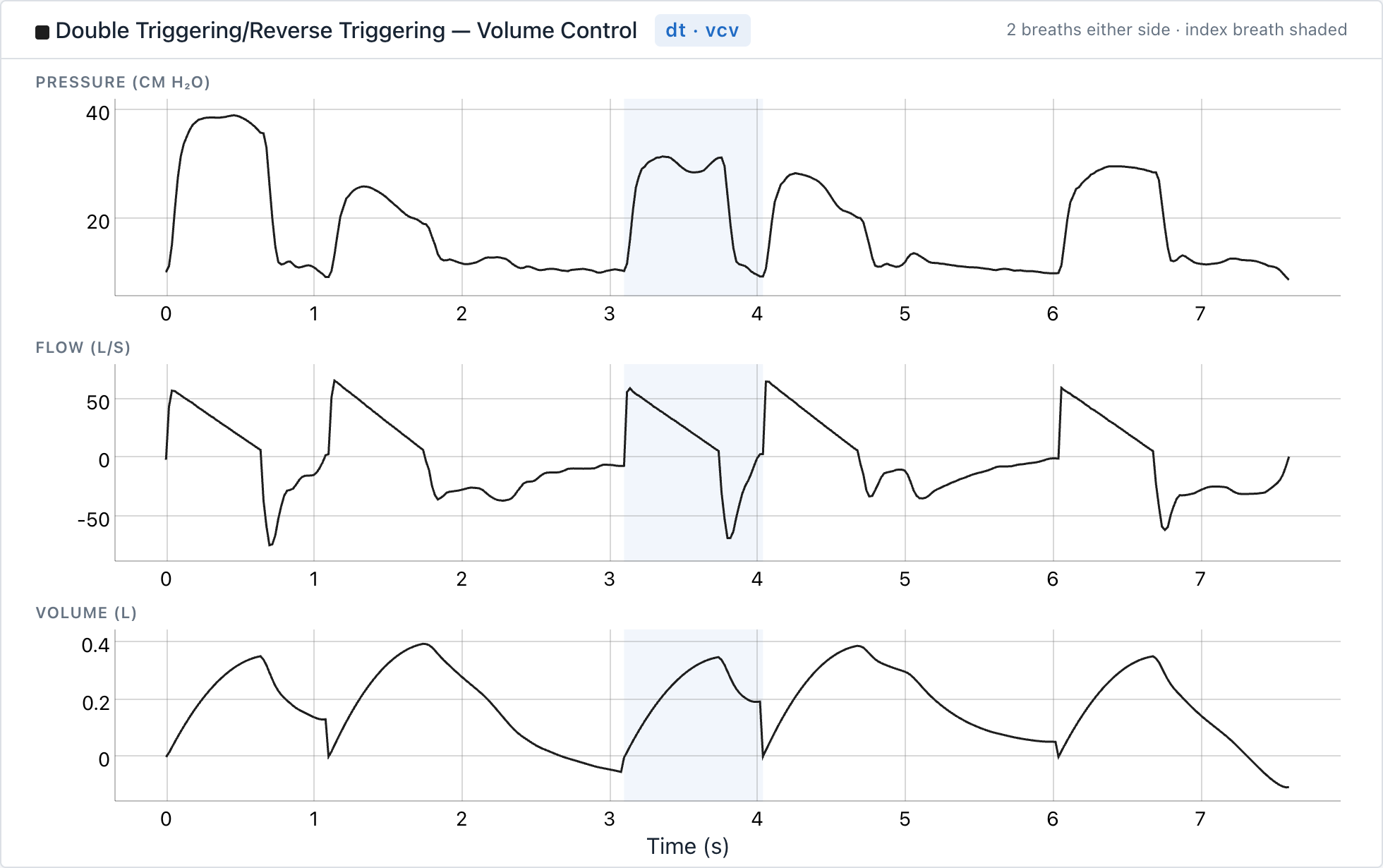 | 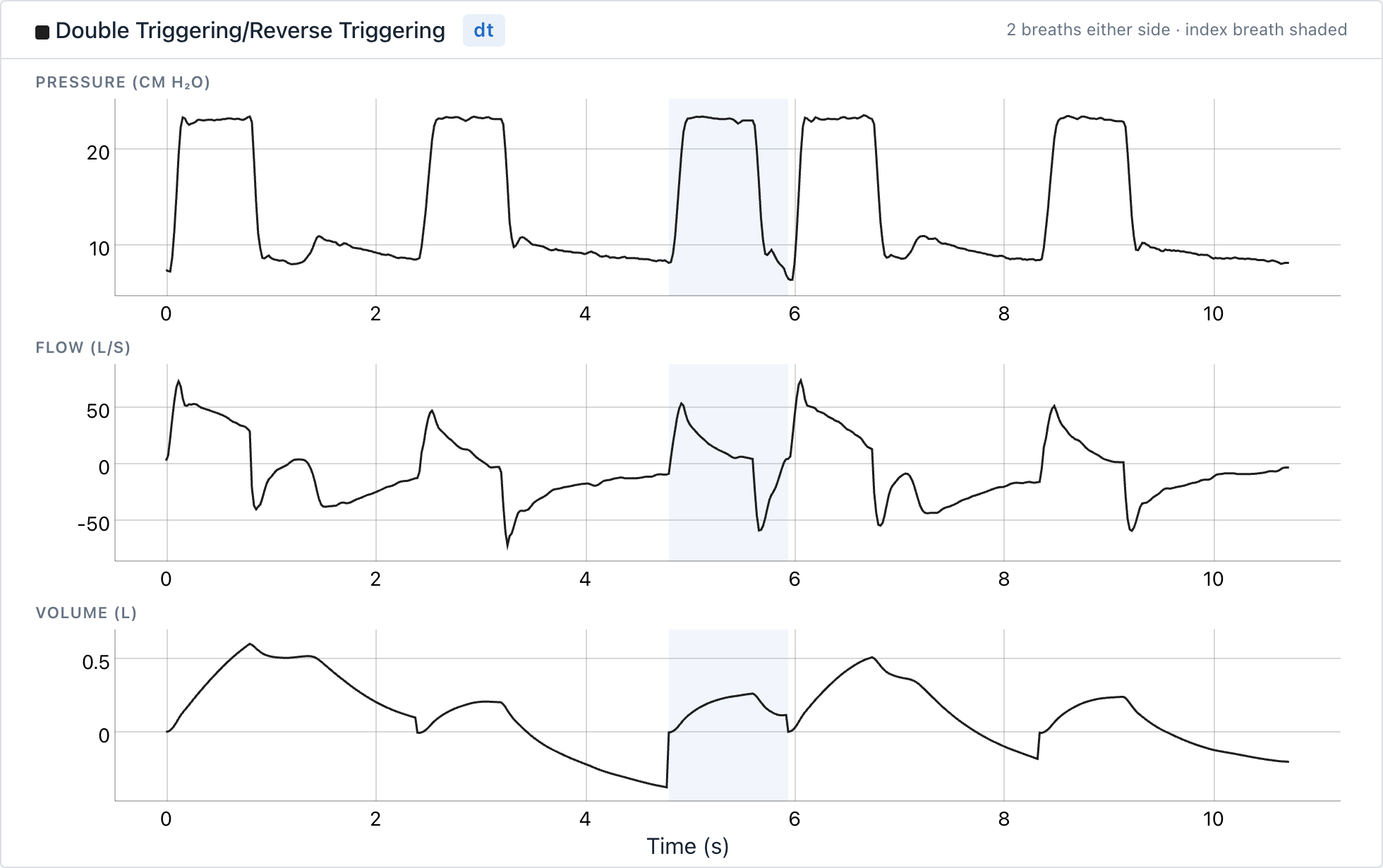 |
