## Supplementary Table S1 for "A Machine Learning Pipeline for Scalable Annotation of Patient-Ventilator Dyssynchrony from Bedside Ventilator Data": Supplementary_Table_S1.docx

**Supplementary Table S1.** Patient–ventilator dyssynchrony (PVD) annotation schema: categories, definitions, and waveform criteria used during expert labeling. The five dyssynchrony subtypes (top five rows) were carried forward as model classes; active exhalation, secretions/cardiogenic oscillations, and unclassifiable were applied during annotation as quality-control or artifact categories and were excluded from model training and evaluation. Normal breaths and the two breath-delivery-type labels (volume- versus pressure-controlled) are model classes but are not waveform-criteria categories and are omitted here. Abbreviations: VC, volume control; PC, pressure control; PEFR, peak expiratory flow rate.

| **Category** | **Definition** | **Annotation criteria** | **Mode applicability** | **Included as model class** |
| --- | --- | --- | --- | --- |
| **Ineffective triggering** | A patient inspiratory effort that fails to trigger a ventilator-delivered breath, such that the effort is not rewarded with mechanical support. | During the expiratory phase, a positive deflection of the flow–time waveform occurs concurrently with a negative deflection of the pressure–time waveform, in the absence of a successfully triggered breath. Both deflections must be visually distinguishable from baseline expiratory flow and pressure variation. | VC and PC | Yes |
| **Flow starvation** | A delivered breath in which ventilator flow and support are insufficient to meet patient inspiratory demand. | In PC modes, in which airway pressure is set by the ventilator, flow starvation manifests as a rounded or convex deformation of the inspiratory limb of the flow–time curve, deviating from the expected natural decay decelerating profile. In VC modes, in which inspiratory flow is set by the ventilator, it manifests as a downward deflection (concavity) of the inspiratory limb of the pressure–time curve, deviating from the expected pressure profile for the set flow. The timing of the deformation may occur in early, mid, or late inspiration. | VC and PC (mode-specific signature) | Yes |
| **Premature cycling** | An asynchrony in which the patient continues inspiratory effort after the ventilator has cycled from inspiration to expiration. | Continued patient inspiratory effort following ventilator cycling produces either a positive deflection of the flow–time curve at the onset of expiration or a blunting of the peak expiratory flow rate. The waveform signature is morphologically similar across VC and PC modes, as the post-cycling flow profile is dominated by the patient's continued inspiratory effort rather than by the ventilator's prior delivery characteristics. | VC and PC | Yes |
| **Delayed cycling** | An asynchrony in which the patient initiates expiration before the ventilator has cycled from inspiration to expiration, typically resulting in expiration against a closed inspiratory valve and an absence of expiratory flow. | In PC mode, the required signature is a positive deflection of the flow–time curve at the end of ventilator-delivered inspiration, indicating the onset of patient expiratory effort against a closed valve. A concurrent upward deflection of the pressure–time curve may be present in severe cases. In VC mode, the required signature is a sharp upward deflection of the pressure–time curve at the end of ventilator-delivered inspiration, reflecting the same physiologic mechanism. | VC and PC (mode-specific signature) | Yes |
| **Breath-stacking dyssynchrony** | Delivery of two consecutive ventilator breaths in rapid succession with insufficient expiratory time between them. This category lumps double triggering and reverse triggering, which cannot be reliably distinguished from single-breath airway pressure and flow waveforms; the mechanistic distinction and its consequences are addressed in the Discussion. | Two consecutive ventilator-delivered breaths separated by a markedly shortened expiratory interval, with the second breath triggered before the flow–time curve returns to baseline. | VC and PC | Yes |
| **Active exhalation** | Recruitment of expiratory muscles during the expiratory phase of a ventilator-delivered breath, producing an augmented expiratory effort beyond passive chest wall and lung recoil. | A convex deformation of the expiratory limb of the flow–time curve, deviating from the expected natural decay profile that characterizes passive expiration. The pressure–time curve is typically unremarkable during expiration, as the rise in intrathoracic pressure produced by expiratory muscle contraction is not transmitted to the airway pressure tracing through an open exhalation valve. | VC and PC | No — excluded from model |
| **Secretions / cardiogenic oscillations** | High-frequency oscillations superimposed on the ventilator waveforms arising from non-dyssynchrony sources: airway secretions producing turbulent flow within the endotracheal tube or large airways, or cardiogenic oscillations from mechanical transmission of cardiac contractions to the airway and thoracic structures. | Rapid, small-amplitude oscillations superimposed on the flow–time curve, the pressure–time curve, or both, not attributable to patient inspiratory or expiratory effort. Secretion-related oscillations are typically coarser, irregular, and most prominent during inspiration or early expiration, and may show a sawtooth appearance of the flow tracing. Cardiogenic oscillations are finer, regular, occur at the patient's heart rate, and are typically most visible during the expiratory plateau when flow is near zero. | VC and PC | No — excluded from model |
| **Unclassifiable** | A quality-control category applied to breaths in which expert classification as either a normal breath or a defined dyssynchrony type was not feasible. | Assigned to breaths in which signal quality limitations, preprocessing artifact, or atypical waveform morphology precluded confident assignment to any of the defined annotation categories. | VC and PC | No — excluded from model |
