## Supplementary Table S2 for "A Machine Learning Pipeline for Scalable Annotation of Patient-Ventilator Dyssynchrony from Bedside Ventilator Data": Supplementary_Table_S2.docx

**Supplementary Methods**

**Decision thresholds**

For each label, the decision threshold maximizing F1 was selected from out-of-fold training predictions by searching the interval [0.10, 0.95] in increments of 0.01. The selected thresholds were 0.296 for normal, 0.483 for volume-controlled ventilation, 0.943 for pressure-controlled ventilation, 0.714 for flow starvation, 0.791 for premature cycling, 0.733 for breath stacking, 0.943 for delayed cycling, and 0.640 for ineffective triggering. Thresholds were held fixed for the supervised baseline and for every semi-supervised round.

**Semi-supervised stopping criteria**

All stopping thresholds were specified a priori. The 0.02 rare-class F1 decrement was chosen to halt expansion as soon as model-assigned label noise began to meaningfully erode performance on the minority subtypes. The 0.001 macro-averaged F1 criterion, applied over four consecutive rounds, was chosen to identify convergence, the point at which additional unlabeled data yielded negligible change in performance.

**Software and computing environment**

Analyses were implemented in Python 3.12.11 using LightGBM version 4.6.0 and scikit-learn version 1.6.1. Breath vectors were stored and queried in PostgreSQL version 15.15 with the pgvector extension. Model training was performed on an NVIDIA DGX Spark with a Grace Blackwell GB10 Superchip (NVIDIA Blackwell GPU with 128 GB unified memory), a 20-core Arm-based NVIDIA Grace central processing unit, and 128 GB LPDDR5X memory.

**Table S2.** LightGBM hyperparameter search space and retained configuration.

| **Parameter (LightGBM name)** | **Values searched** | **Retained value** |
| --- | --- | --- |
| Number of estimators (*n_estimators*) | 300, 500, 800 | 500 |
| Learning rate (*learning_rate*) | 0.02, 0.05, 0.1 | 0.1 |
| Number of leaves (*num_leaves*) | 31, 63, 127, 255 | 127 |
| Minimum child samples (*min_child_samples*) | 20, 50, 100 | 50 |
| Subsample ratio (*subsample*) | 0.7, 0.8, 1.0 | 0.7 |
| Column subsample ratio (*colsample_bytree*) | 0.7, 0.8, 1.0 | 0.7 |
| L1 regularization (*reg_alpha*) | 0, 0.1, 1.0 | 0 |
| L2 regularization (*reg_lambda*) | 0, 0.1, 1.0 | 0.1 |

Twenty-five configurations were sampled at random from the 8,748 possible combinations and evaluated on a 150,000-breath development subsample drawn from the training and development partition. Selection was by macro-averaged area under the precision-recall curve under 3-fold cross-validation, averaged across labels with at least one positive example in the fold. The held-out test set was excluded from all tuning. Class weighting (class_weight = balanced) was fixed across all configurations and was not tuned. The retained configuration was used without further modification for the supervised baseline and for every semi-supervised round. Random seed: 42. Bagging frequency (subsample_freq): 1.
